# A Tale of Two Waves: Diverse Genomic and Transmission Landscapes Over 15 Months of the COVID-19 Pandemic in Pune, India

**DOI:** 10.1101/2022.11.05.22281203

**Authors:** Divya Niveditha, Soumen Khan, Ajinkya Khilari, Sanica Nadkarni, Unnati Bhalerao, Pradnya Kadam, Ritu Yadav, Jugal B Kanekar, Nikita Shah, Bhagyashree Likhitkar, Rutuja Sawant, Shikha Thakur, Manisha Tupekar, Dhriti Nagar, Anjani G. Rao, Rutuja Jagtap, Shraddha Jogi, Madhuri Belekar, Maitryee Pathak, Priyanki Shah, Shatakshi Ranade, Nikhil Phadke, Rashmita Das, Suvarna Joshi, Rajesh Karyakarte, Aurnab Ghose, Narendra Kadoo, LS Shashidhara, Joy Merwin Monteiro, Dhanasekaran Shanmugam, Anu Raghunathan, Krishanpal Karmodiya

**Author notes:** Equal contributing authors (Divya Niveditha, Soumen Khan).

## Abstract

The modern response to pandemics, critical for effective public health measures, is shaped by the availability and integration of diverse epidemiological outbreak data. Genomic surveillance has come to the forefront during the coronavirus disease 2019 (COVID-19) pandemic at both local and global scales to identify variants of concern. Tracking variants of concern (VOC) is integral to understanding the evolution of severe acute respiratory syndrome coronavirus 2 (SARS-CoV-2) in space and time. Combining phylogenetics with epidemiological data like case incidence, spatial spread, and transmission dynamics generates actionable information. Here we discuss the genome surveillance done in Pune, India, through sequencing 10,496 samples from infected individuals and integrating them with multiple heterogeneous outbreak data. The rise and fall of VOCs along with shifting transmission dynamics in the time interval of December 2020 to March 2022 was identified. Population-based estimates of the proportion of circulating variants indicated the second and third peak of infection in Pune to be driven by VOCs Kappa (B.1.617.1), Delta (B.1.617.2), and Omicron (B.1.1.529) respectively. Integrating single nucleotide polymorphism changes across all sequenced genomes identified C (Cytosine) > T (Thymine) and G (Guanine) > T (Thymine) substitutions to dominate with higher rates of adaptive evolution in Spike (S), RNA-dependent RNA polymerase (RdRp), and Nucleocapsid (N) genes. Spike Protein mutational profiling during and pre-Omicron VOCs indicated differential rank ordering of high-frequency mutations in specific domains that increased the charge and binding properties of the protein. Time-resolved phylogenetic analysis of Omicron sub-lineages identified specific recombinant X lineages, XZ, XQ, and XM. BA.1 from Pune was found to be highly divergent by global sequence alignment and hierarchical clustering. Our “band of five” outbreak data analytics that includes the integration of five heterogeneous data types indicates that a strong surveillance system with comprehensive high-quality metadata was critical to understand the spatiotemporal evolution of the SARS-CoV-2 genome in Pune. We anticipate the use of such integrated workflows to be critical for pandemic preparedness in the future.

---

At the leading edge of the SARS- CoV-2 pandemic are emerging Variants of Concern (VOC) and Variants of Interest (VOI) of the virus, driving global infection. The diversity of such variants and emerging lineages ^1–5^ potentially shapes transmissibility, disease severity, and immune escape mechanisms that ultimately impact public health. Tracking the emergence and spread of SARS-CoV-2 lineages ^1–6^ using phylogenetics has been critical to informing the timing and stringency of COVID-19 public health interventions ^7–11^. The first known case that spiraled India into the COVID-19 national pandemic was reported in January 2020 ^12^. The national infection case toll is reaching 45 million people with the death toll reaching 0.5 million ^13^. Pune, a city with a 7 million population, recorded its first COVID-19 case in March 2020, with a traveler flying in from Dubai^14^. Pune has recorded 1.1 million infection cases ever since with the death toll reaching 20,000 ^15^

In this study, a city-wide network of researchers, clinicians, and pathology diagnostic laboratories was formed to investigate the changing landscape of the genome of SARS-CoV-2 infection and its correlation to the COVID-19 pandemic during the time frame spanning December 2020 to March 2022 in Pune, India. Genome surveillance of 10,496 SARS-CoV-2 infected patients was key to the timely identification of SARs-CoV2 variants that impacted public health decision-making by local authorities in Pune. As a modern informed response to the pandemic, a “band of five” outbreak data analytics approach ^16^ was used. This integrated the genomic data (Band 1) of the virus through molecular phylogenetics with key outbreak data including sample collection dates and case numbers (Band 2), demographics like age and gender (Band 3-4), and geospatial mapping (Band 5). The population-based proportion of circulating variants across time identified the VOCs driving infection peaks. Mutations in Spike protein that were potentially selected shaping higher binding and transmission rates in Omicron VOC were identified in Pune. Hierarchical clustering analysis identified conserved and unique key residues that impacted the infection rates. We foresee the utility of such scalable and integrated outbreak analytics workflows to not only understand the evolution of the virus and the pandemic but drive public health decisions and policy making.

Next-generation sequencing was used to sequence 10,496 SARS-CoV-2 genomes prepared from COVID-19 positive (by RT-PCR) patients (Supplemental data; See Methods for details). Oxford Nanopore Technologies was used to generate sequence data (n=2868) wherever low throughput sequencing was required including outbreak clusters, re-infection, and vaccine breakthroughs, while Illumina NextSeq550 platform (n= 7588) was used to generate high throughput data from COVID-19 positive samples in the Pune region. Variant analysis of circulating SARS-CoV-2 genomes was performed to understand the accumulation of mutations ^17^, a fundamental driving force for viral evolution within Pune. The lineage distribution across the time span of 15 months from December 2020–March 2022 showed a few striking trends. Phylogenetic analysis (See Methods for details) identified 182 SARs-CoV2 lineages from the sequenced samples (Fig 1a), out of which 27 were variants of concern (VOCs) as identified by WHO (Supplemental data table).

**Figure 1:**
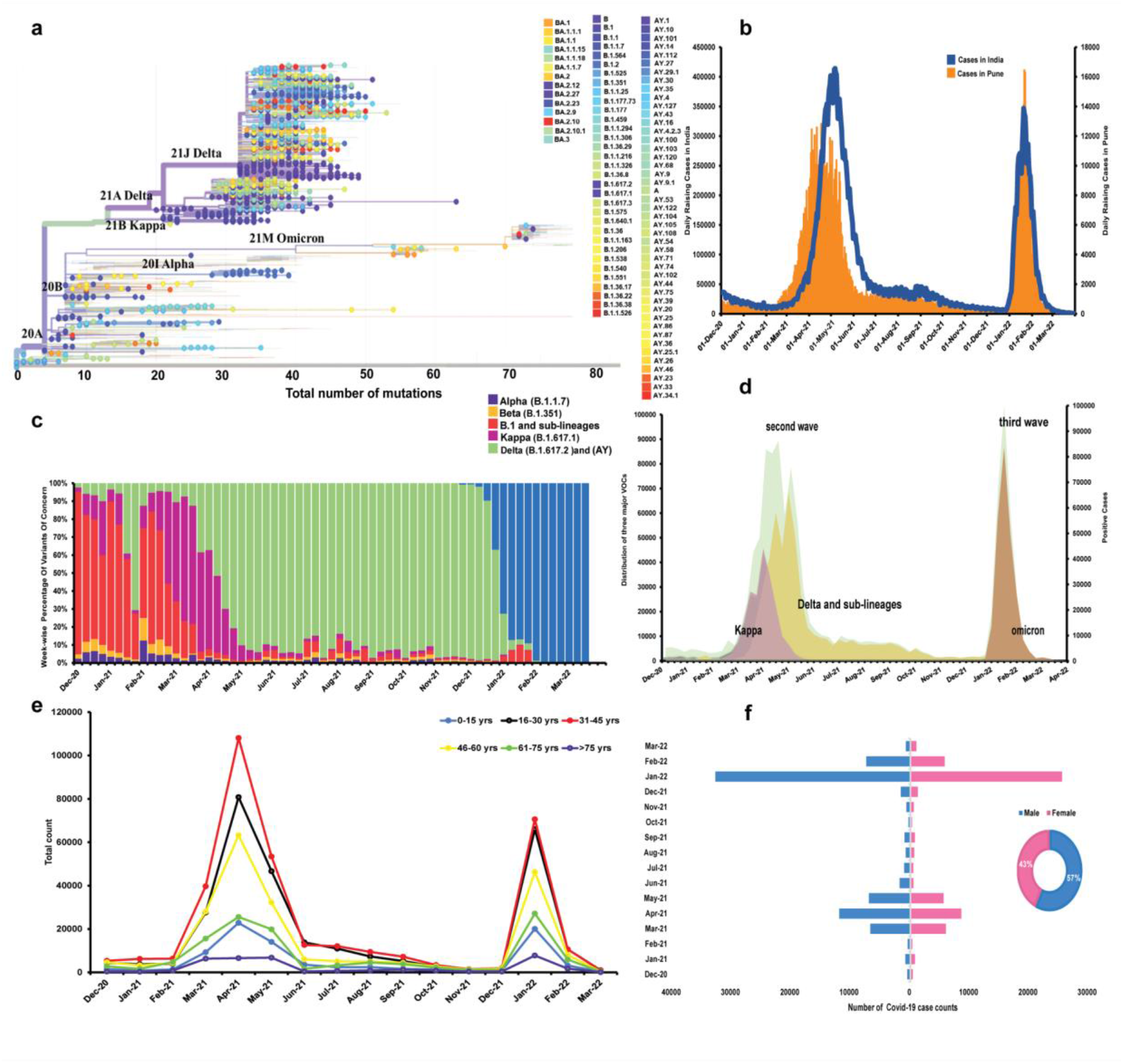
Epidemiological profile of SARS-CoV-2 for the time interval of December 2020 to March 2022 for Pune. **a)** Phylogenetic relationship tree of 182 lineages of SARS-CoV-2 detected in Pune. **b)** Distribution of Daily Positive case numbers in Pune compared to India. **c)**Percentage Proportion of VOCs seen in the Pune population through the course of the time interval. **d)** Dynamic landscape of the VOCs, Kappa, Delta, and Omicron, predominantly driving the Second and Third-wave for Pune. **e)** Age distribution indicating infectivity of SARS-CoV-2 in Pune. **f)** Gender distribution indicating infectivity of SARS-CoV-2 in Pune.

Epidemiological surveillance data gives transmission context to the identified variants ^18^. The incidence of COVID-19 within Pune along with that in India during the period spanning dates for the time interval of December 2020 to March 2022 is shown in (Fig 1b) along with the case incidence time series data as a week-wise histogram. This clearly indicates the time span of the second wave was from March 2021 to June 2021 although there were some cases beyond June. The time span of the third wave was from December 2021 to March 2022 in Pune. The transmission dynamics reflect (Fig 1c) the rise and fall of the VOCs through the course of the pandemic which started off with the Kappa (B.1.617.1) variant driving it. However, Delta (B.1.617.2) and its sub-lineages became predominant later during the second wave as the infections peaked, and Omicron (B.1.1.529) sub-lineages dominated the infections in the third wave. Although Delta was detected early on in December 2020, the prevalent lineages included B1 sub-lineages, Alpha, and Beta followed by Kappa until mid-April 2021 before Delta and sub-lineages became the predominant VOCs in circulation. Delta and sub-lineages were the dominant and prevalent lineages until early December 2021 in the Indian population.

Three weeks after the detection of VOC Omicron in South Africa, the first Omicron case was sequenced ^19–21^ and reported in Pune in December 2021. With an infection rate reported to be high as compared to Delta, the Omicron VOC was the only predominant VOC in circulation by the end of 2021 ^22^. The prevalence of Omicron in samples sequenced from Pune was close to 80% by the third week of January. While sub-lineages of Delta were detected in 5% of the samples sequenced when Omicron emerged in December, B.1.617.2 was detected at frequencies less than 1% in January.

The sampling strategy (for details refer to methods section) for both retrospective and real-time surveillance was designed to accurately track lineage transitions and the rate of change of prevalence of variants over time for both low (5%) and high (70%) prevalence. Such a sampling strategy allowed statistically robust estimates of the prevalence of VOIs and VOCs, especially when the number of identified cases was high. Such methods allow the generation of population-based estimates of the proportion of each circulating variant among all SARS-CoV-2 infections ^23^.

To account for the varying proportion of samples sequenced over time in relation to the infected population, the variants were scaled to the infected cases reported in Pune (Fig 1d). The peaks of the scaled proportions of VOCs Delta and Omicron matched the peaks of infection during the second and third waves. Kappa, Delta, and its sub-lineages remained the only VOCs circulating at a high proportion from March–November 2021 but were rapidly replaced by Omicron in mid-December 2021. The rise of the Kappa, Delta, and Omicron variants was associated with major surges in COVID-19 cases during March–July 2021 and December 2021–January 2022. The resurgence of transmission across Pune (Fig 1b) coupled with the dynamic landscape of prevalent VOCs in 2021 underscores the importance of data analytics pipelines and robust genomic surveillance.

The fifth band of data relating to demographics of age indicated more infections in the age group of 31-45 years when the prevalent VOC was Delta and it shifted to the age group of 16-30 years when the prevalent VOC was Omicron (Fig 1e). We observed that the infectivity rate is higher in men than in women. However, gender did not seem to play much of a major role in the infected case numbers based on the monthly and overall distribution of infected cases (Fig 1f).

Phylogeographic distribution or geospatial dynamics was studied for the lineages sequenced ward-wise in Pune city (Fig 2b). Wards are administrative subdivisions of Pune city (Fig 2a). The ward-wise rise of lineages potentially suggests specific patterns of migration and travel around Pune. High and medium population density wards (KB, DS, HM, AB) were equally affected in the peaks of infection driven by both Delta and Omicron VOC. The ward DP that was highly infected based on the city COVID dashboard data during the second wave, showed very less infectivity in the third wave by Omicron and this could be due to increased immunity from Delta infections. The flow of infections across time in 2022 is observed to be HM to DP road indicating potential travel flow in this direction. Further characterization of travel patterns would allow another band of data to be integrated into these pipelines to understand the directionality of infection.

**Figure 2:**
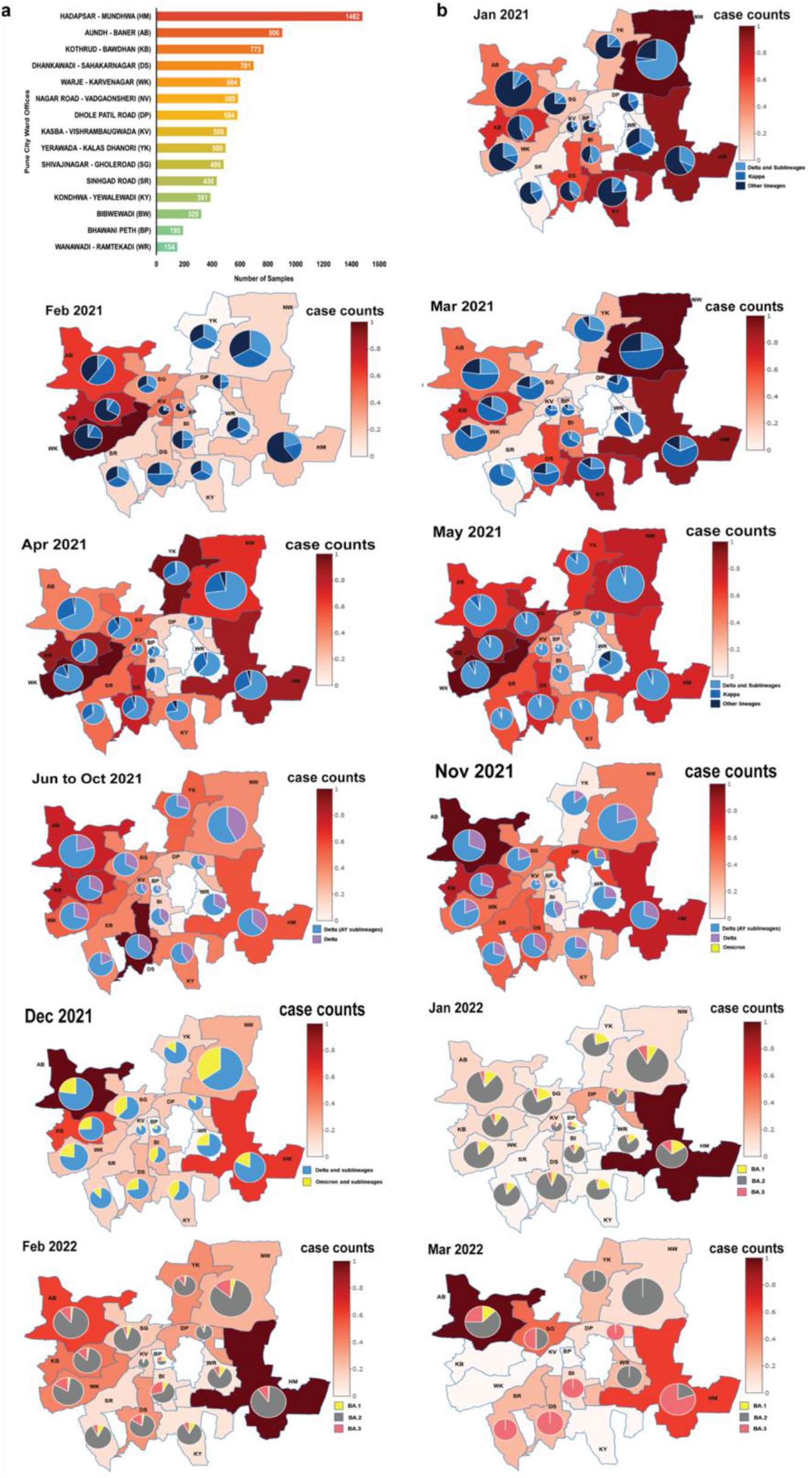
Transmission dynamics of SARS-CoV2 lineages in Pune samples. **a)** The bar plot shows the occurrence of positive cases during the second and third waves in the wards of Pune city. **b)** The choropleth maps of the wards represent the positive samples collected and sequenced from Jan 2021 to Mar 2022 and the pie charts within the choropleth maps show the distribution lineages identified from the sequencing data.

The distribution of mutations on the complete coding region of SARS-CoV-2 of 10,496 sequenced samples is represented (Fig 3a) in genomic order by gene and position on the genome (x-axis) and the frequency of its occurrence on the y-axis. The number of unique mutations (NU) is determined by counting the same type of mutation in different genome isolates only once, while the number of non-unique mutations (NNU) is calculated by counting the same type of mutation in different genome isolates repeatedly ^24^. The unique and non-unique mutations can be classified as substitutions (transversions & transitions), deletions, and frameshifts. The Manhattan plot depicts around 890 mutations that are unique and hence have been identified only once and 9101 common ones along with their mutation occurrence frequency. The common mutations with higher frequency form a pattern of dots higher on the y-axis like a skyline. The genes are represented on the x-axis as they are sequentially positioned on the SARS-CoV-2 genome.

**Figure 3:**
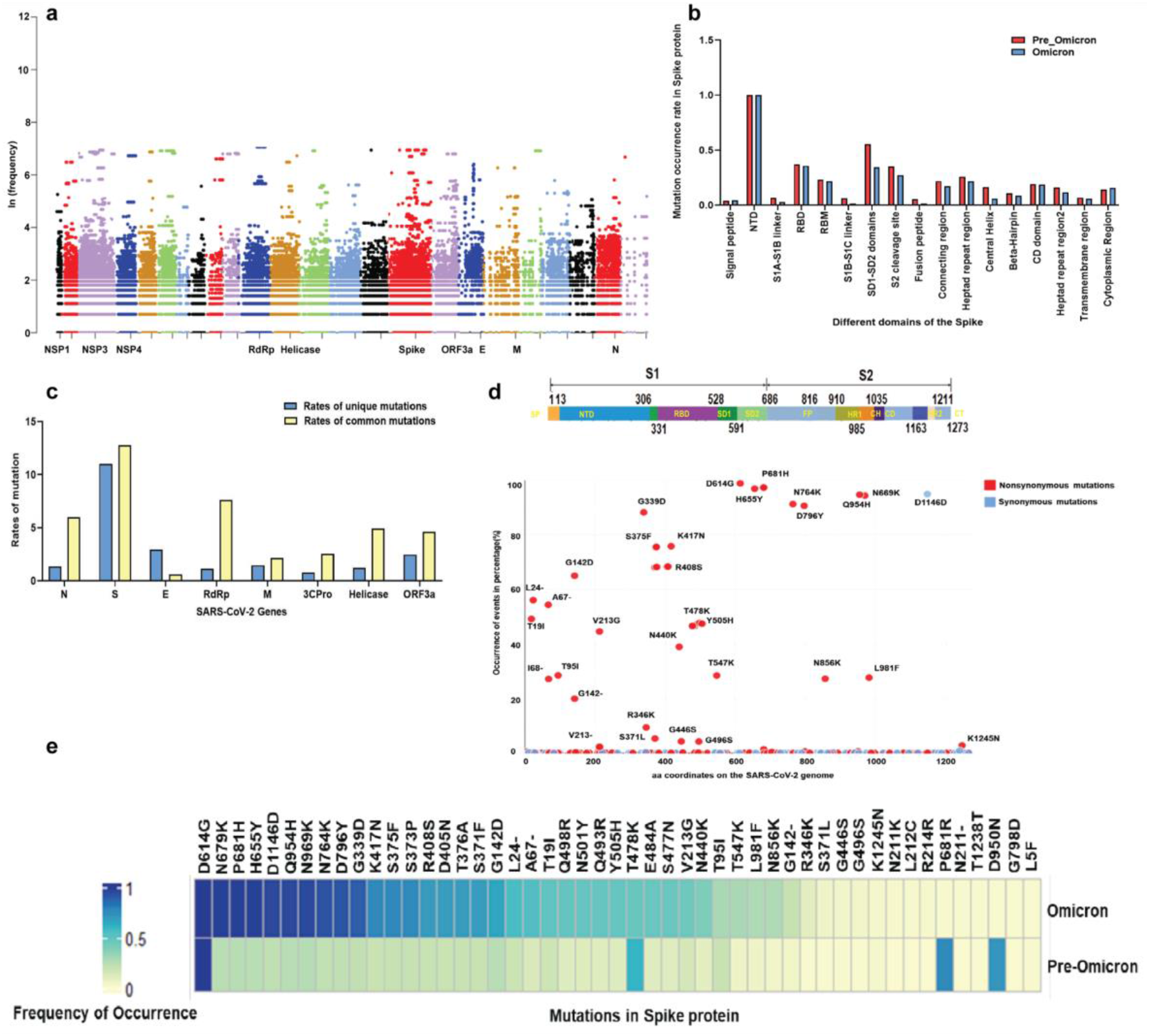
Mutation profiling of SARS-CoV2 genome of Pune samples. **a)** The Manhattan plot shows the distribution of mutations across 26 proteins of the SARS-CoV2 genome. The y-axis represents the natural log frequency for each mutation present at a specific position on the genome. Only a few key genes of SARS-CoV2 (protein) names are labeled **b)** The bar graph represents the comparison of the rate of mutation occurrence of mutations between non-omicron vs omicron samples **c)** The bar plot represents the rate of unique and common mutations and their occurrence in different genes within the genome of SARS-CoV2. **d)** The common interface graph is showing the mutation frequency in SARS-CoV2 Spike protein, with the occurrence of each mutation represented on the y-axis and its corresponding protein coordinate on the x-axis. The red dots indicate an amino acid changing “Nonsynonymous” mutations and the blue indicates “Synonymous” mutations. **e)** The frequency heatmap depicts the occurrence rate of the top 50 mutations in non-omicron sequences during the second wave and the occurrence rate of the same mutations in omicron sequences during the third wave.

Among the 10,496 complete genome sequences, 98 unique single mutations were detected on the S protein. The distribution of 12 Single Nucleotide Polymorphisms (SNP) types among unique and non-unique mutations and the rate of mutation (Table 1) on the S protein of SARS-CoV-2 in Pune indicate positive selection for certain genes (Table 2, Supplemental data). The overall rate of mutations for the common unique or conserved mutations (Rc) is highest for spike (S: 12.7%) followed by RNA-dependent RNA polymerase (RdRp: 7.5%) and the Nucleocapsid (N:6%) protein as expected (Fig 3c). The rate of mutations for the unique mutations (Ru) is highest for Spike (S:11%) followed by the Envelope protein (E: 3%). The Envelope protein is the only structural protein that is less convergent in mutation (Fig 3c) indicating slower evolution. The two dominant substitutions and SNP types across the genome and in Spike are C > T and G > T (Table 1). This could be potentially due to the innate host immune response via Apolipoprotein B mRNA editing enzyme, catalytic polypeptide (APOBEC), and Adenosine deaminase acting on RNA (ADAR) gene editing ^25–27^. Adaptive evolution rates suggest positive selection for the Spike, E, and Orf3a genes and purifying selection for Helicase and RdRp genes (Table 2, Supplemental data) ^28^

**Table 1:**
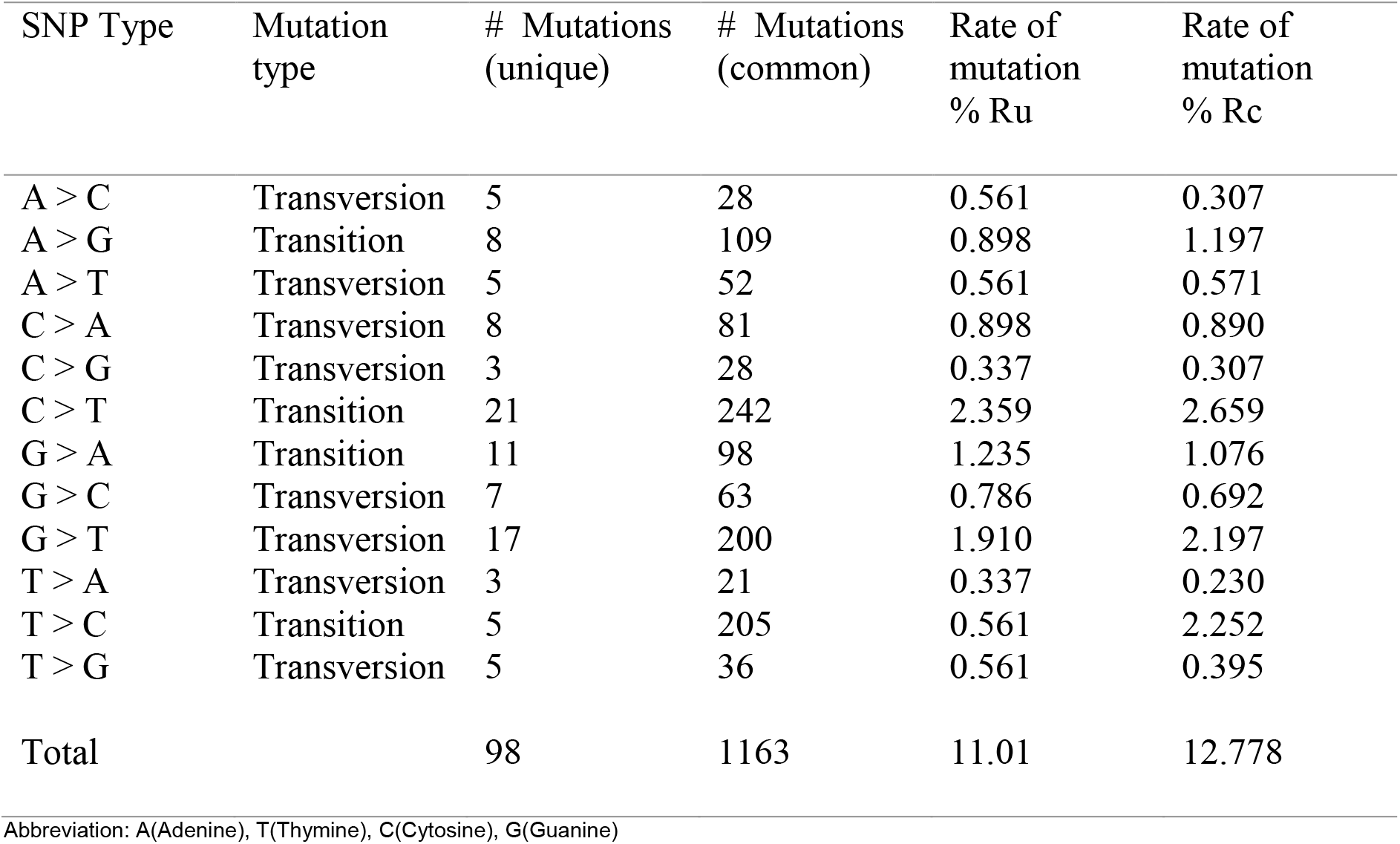
Classification and Distribution of Single Nucleotide Polymorphisms across samples sequenced.

| SNP Type | Mutation type | # Mutations (unique) | # Mutations (common) | Rate of mutation % Ru | Rate of mutation % Rc |
| --- | --- | --- | --- | --- | --- |
| A > C | Transversion | 5 | 28 | 0.561 | 0.307 |
| A > G | Transition | 8 | 109 | 0.898 | 1.197 |
| A > T | Transversion | 5 | 52 | 0.561 | 0.571 |
| C > A | Transversion | 8 | 81 | 0.898 | 0.890 |
| C > G | Transversion | 3 | 28 | 0.337 | 0.307 |
| C > T | Transition | 21 | 242 | 2.359 | 2.659 |
| G > A | Transition | 11 | 98 | 1.235 | 1.076 |
| G > C | Transversion | 7 | 63 | 0.786 | 0.692 |
| G > T | Transversion | 17 | 200 | 1.910 | 2.197 |
| T > A | Transversion | 3 | 21 | 0.337 | 0.230 |
| T > C | Transition | 5 | 205 | 0.561 | 2.252 |
| T > G | Transversion | 5 | 36 | 0.561 | 0.395 |
| Total |  | 98 | 1163 | 11.01 | 12.778 |

Abbreviation: A(Adenine), T(Thymine), C(Cytosine), G(Guanine)

Spike protein that facilitates virus entry into the host cells is a trimeric glycoprotein, that binds to the ‘angiotensin-converting enzyme 2’ (ACE2) surface receptors of the host ^25,29^. Most antibodies neutralize coronavirus infection by binding to the receptor-binding subdomain of S1 in Spike and block ACE2 receptor interactions. Mutations in different domains within the Spike gene in pre-omicron and omicron sequences have been delineated (Fig 3b). Thus, the high numbers of mutations in immunodominant regions including the receptor binding domain (RBD and the receptor binding motif (RBM) of the Spike (S) gene (Fig 3b, d) of VOC Omicron potentially have an adverse effect on antigenicity ^30,31^ (Supplemental data).

The S gene from the VOC Omicron was thus mapped on the structural domains of the spike ^24,32^ to identify a similar frequency of mutation events in all domains including the RBD and RBM region of the S gene. (Fig 3e). On analysis of the mutation occurrences on the S gene for the Omicron VOC, several high-frequency SNPs have been identified that increase the number of charged residues that potentially would impact electrostatic attraction during host-virus interaction. Binding free energy (BFE) changes (Supplemental data) were extracted from Chen J et al for all 46 non-degenerate mutations in the S protein and human ACE2 interactions sites ^25^. 19 high frequency (n>100) single-site non-degenerate mutations in the S1A and S1B domains of the S protein in the Omicron VOC impacting BFE were identified. 78% of these showed a positive BFE change indicating the strengthening of the binding between S protein and ACE2, giving rise to potentially more infectious variants. The rest potentially weaken the binding between them and could play a role in immune escape ^25,30^.

The mutations observed in the RBD and the S1/S2-domain of the Omicron variant (Fig. 3d) increased the number of positively charged amino acids by 9 in Omicron compared to an increase in charge by 4 in the Delta variant. Some of the new positive charges in the S1/S2 domain compared to the wild-type ^5^ led to an amplification of the Cardin-Weintraub motif (CWM; a peptide series of hydrophobic and cationic AAs) on the surface of the spike protein involved in binding to the host ACE protein ^33^.

The CWM in wt SARS-CoV-2 corresponds to amino acid sequences of ‘XBBXBX’ and changes to ‘XBBBXBX’ in the VOC Delta via the cationic patch L452R and T478K. In the VOC Omicron, the cationic patch further expands by Q493R, Q498R, and Y501H on the RBM region which promotes even stronger binding with the heparan sulfate proteoglycans involved in host cell entry. Several studies have shown that mutations in the RBD, mutations in the RBM such as K417, S477, T478, E484, and N501 also seen in the Omicron variant potentially enhance binding to ACE2 ^31,33^. A change of residue charge due to mutation potentially increases the resistance of the virus to vaccine-elicited antibodies ^31^. Coupled with the BFE change may also decrease antibody recognition and promote immune escape ^25,34^. The mutational frequency of SNPs in VOC Omicron is higher for most AA changes except for 3 AA residues (T478K, P681R, D950N). These SNPs are lost during the adaptive evolution of the virus as seen in legacy data ^32^.

The distribution of Omicron lineage sub-variants and their rise and fall from mid-December until the end of March 2022 (Fig 1d) indicate their high prevalence. Omicron was comprised of three sister lineages: BA.1, BA.2, and BA.3 (Fig 4a). The sub-lineage BA.1 was the first lineage detected in Pune and was slowly taken over completely by BA.2. There was a simultaneous but slow rise of BA.3. Bayesian phylogenetic methods revealed that the three lineages were distinct from each other. Three recombinant variants were identified belonging to the X series in Pune. Originally discovered in Brazil, XQ recombinant was a lot like BA.1. XM that was first discovered in the Netherlands was more like BA3 ^35^ and a heretofore globally undiscovered XZ showed a high resemblance to BA.2. Certain recombinant lineages that belong to the X series that were first identified in the UK first were also detected ^35^. Recombinant variants are hybrid products of different types of the SARS-CoV-2 variants that were either previously circulating or actively circulating now through the exchange of genetic material between the viruses and the host cells ^35^.

**Figure 4:**
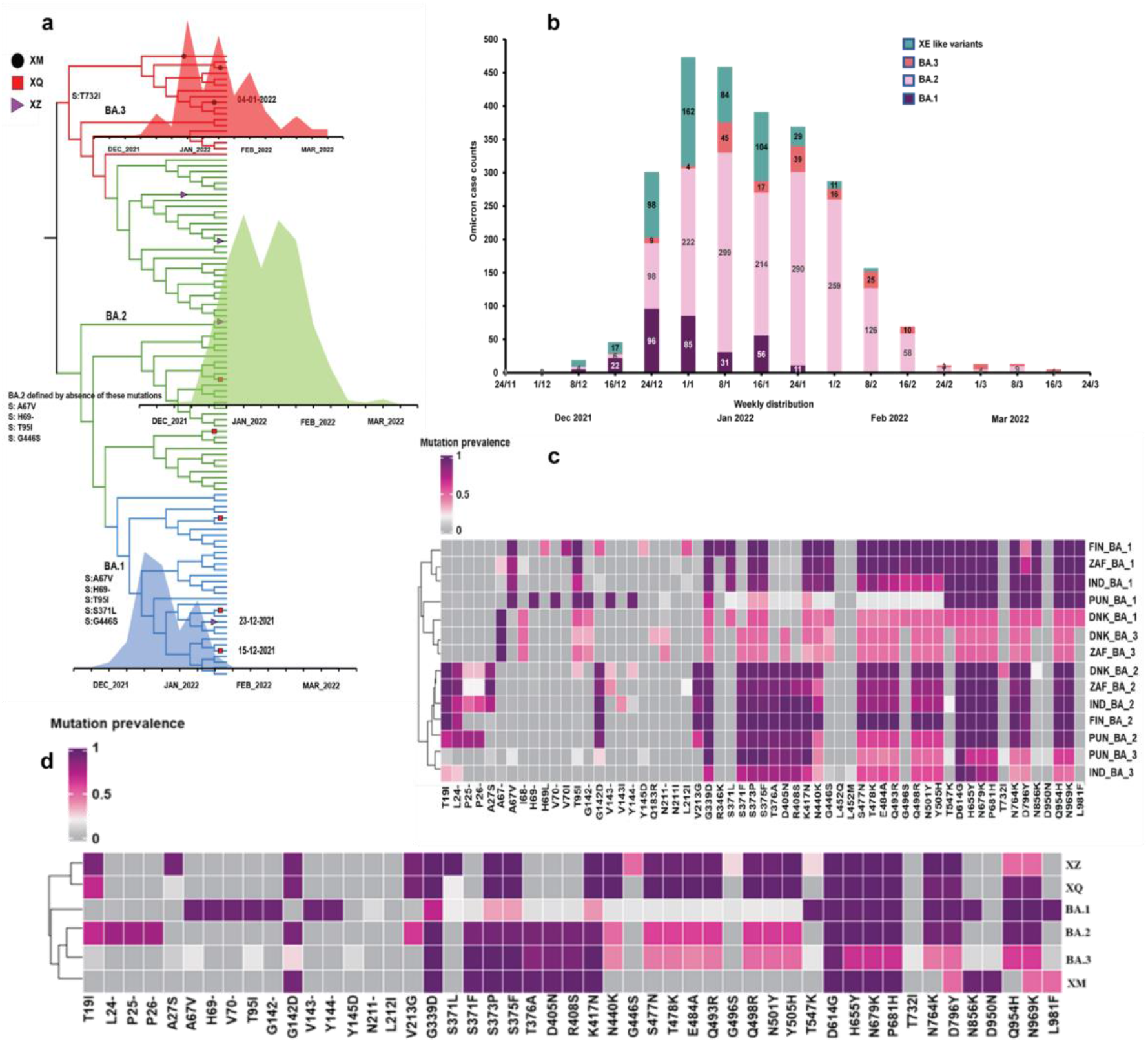
The Mutation profiling of Spike protein in Omicron sequences. **a)** The time-resolved beast plot illustrates the emergence and spread of three variants of omicrons (BA.1, BA.2, and BA.3) during the third wave in Pune. The spatiotemporal reconstruction of the spread of BA.1 is indicated in blue while for BA.2 and BA.3 it is indicated in green and red. The square, triangle, and circle at the branch nodes represent the emergence of the recombinants (XQ, XZ, and XM) according to their inferred time of occurrence. The area plots show the distribution of BA.1, BA.2, and BA.3. The mutation which defines each clade has been labeled. **b)** The bar graph represents the distribution of each omicron variant (BA.1, BA.2, and BA.3) along with the emergence and spread of recombinants (XQ, XZ, and XM). BA.1 is shown in maroon whereas BA.2 and BA.3 are shown in pink and orange and all the recombinants are represented in green **c)** The heatmap showcases the diversity in the mutation profile of omicron at the global level **d)** The clustergram heatmap of omicrons of Pune samples showcases the mutation profile of the recombinants clustering together with BA.1, BA.2 and BA.3.

The mutational profiles of the spike protein in BA.1, BA.2, BA.3, and X variants have both convergent and unique profiles ^28,36,37^ and the sequences were analyzed for features to understand the drivers for the replacement of the BA.1 variant by BA.2 in the population (Fig. 4b). The Spike protein sequences from 35,827 sequenced samples across the world were analyzed using Multiple Sequence Alignment algorithms and hierarchical clustering. The three prominent Omicron sub-lineage (BA.1, BA.2, and BA.3) sequences from Pune (2540) were compared with sequences from India (7935), South Africa (9484) and Denmark (8979), Finland (6346) in the time interval ranging from December 2021 to March 2022. The prevalence of the spike gene mutations across the three prominent Omicron sub-lineage sequences in Pune (BA.1, BA.2 and BA.3) identified a differential prevalence of the common lineage-defining mutations (Fig 4c) across the lineages. The BA.1 VOC in Pune was the most divergent when compared to the global Omicron sequences. The prevalence of the common mutations from AA residue 346 to 505 was very low. BA.2 and BA.3 VOCs with sequence conservation within the global sequence set had an increasingly high prevalence of mutations in the N-Terminal Domain (NTD) of the spike protein. The lower prevalence in BA.3 of the mutations V213G and T478K indicate a lower fitness advantage also correlating to the eventual loss of T478K. Prevalence of the lineage-defining mutations in Pune showcased reversed prevalence of G142- and G142D amongst BA.1 and BA.2. *In silico* analysis of the G142D mutation suggests a disturbance of the surface topography of the ‘super site’ epitope that binds NTD-directed neutralizing antibodies (NAbs) ^38–40^. The changing pattern for G142 to D142 or G142- and V213 to G213 or G213-observed at multiple time points and across Delta VOC sub-lineages indicates the rapid generation of multiple variants for selection. Although the etiology of these recurrent mutations is unclear, it could potentially result from host-cell induced RNA editing for the emergence and or re-emergence of mutations for the selection of more biologically ‘fit’ genotypes^25–27^.

The heatmap (Fig 4c) further highlights the differential distribution of the lineage-defining mutations across the different geographical locations. For example, in the sub-lineage BA.1, deletion mutations such as S:H69- and S: V70-, S: G142-, S: V143-, S: Y144-and SNPs like T95I were highly prevalent and drop out in BA.2. However, at residue 142 an SNP, S: G142D was selected for and is persistent in BA.3 The mutations around AA143-145 are variable (S: V143-S: Y144-S: Y145D) present in Pune and India and based on timelines, travel patterns could potentially be the cause of transmission. The delta lineage defining D950N is completely lost in Omicron. Loss and gain of mutations could potentially indicate the gain of a fitness advantage through viral adaptation to evolving host immunity between the sub-lineages.

Hierarchical clustering analysis (Fig 4d) of mutation profiles of the spike protein of the samples sequenced in Pune alone shows a few remarkable features including the divergence and convergence of X recombinants. The very first recombinants XQ and XZ were more like BA.1 and first emerged during the 2nd and 3rd week of December 2021. Sharing conserved AA residues with BA.2 they continue to be present in February 2022 when BA.2 was highly prevalent in Pune. Recombinant XM is closer to BA.3 in many sequence features but seems to have retained the delta-defining D950N SNP that was not present in the other sub-lineages of Omicron. In the AA region 339-505, some lineage-defining mutations decreased in prevalence in XM and increased in prevalence in XQ and XZ indicating a potential host editing for a fitness advantage. A mutation like T732I has been observed in BA.3 for the first time that with the neighboring two amino acids favoring β-sheet formation vis a vis coil formation ^40^. This would potentially have an impact on the conformation and functions of the spike protein.

## Conclusions

The SARS-CoV-2 virus continues to evolve. Taken together, our “band of five” outbreak data analytics indicates that a strong surveillance system locally along with methods for integration of comprehensive high-quality meta-data was critical to understand the spatiotemporal evolution of the SARS-CoV-2 genome. Our study highlights the molecular epidemiology trends of SARS-CoV-2 in Pune in the time frame December 2020 to March 2022. Integration of the phylogenetic data (VOC/VOI) with heterogeneous epidemiological data tracked the VOC that was driving up the infection cases in Pune and the geospatial spread of the virus in the city. The rates of evolution of different substitutions in the genome of SARS-CoV-2 underscored the significance of specific amino acids in the spike protein. Detailed molecular analysis coupled with legacy data on mutational impact predicted the potential impact on binding to host receptors and immune system escape. We foresee such pipelines for integrated analysis of SARS-CoV-2 genome surveillance data and epidemiological outbreak data as critical to tracking and predicting future waves of the pandemic.

## Materials and Methods

### 1. Sample collection and sequencing of patient samples

Individuals (n=10456) who were found to be RT-PCR positive for SARS-CoV-2, were sequenced to determine the lineage of the causative virus. Oxford Nanopore sequencing was undertaken for 2686 samples and the Illumina NextSeq550 platform for 7588 samples.

### 2. Sampling strategies for sequencing samples across Pune

Sampling strategies for both retrospective and real-time surveillance were narrowed down in order to be able to accurately track lineage transitions and rate of change of prevalence over time for both low (5%) and high (70%) prevalence with reasonable precision. Sequencing was performed for 300-400 samples every two weeks which allowed for estimating 5% prevalence at +-2% precision and 70% prevalence at +-5% precision, with a p-value >= 0.95. Minimum essential metadata associated with each sample was collected which includes patient-ID, ICMR_ID, address, Age, Gender, Location, date, and time of sampling. Although symptoms during testing, vaccination status, hospitalization status, and disease severity were not always available.

### 3. SARS-CoV-2 whole genome sequencing using Oxford Nanopore Technology (ONT)

For cDNA synthesis, 100 ng total RNA from the swab samples was taken for first strand synthesis using Superscript IV (ThermoFisher Scientific, Cat. No. 18091050) or Luna Script RT Master mix (New England Biolabs). cDNA thus obtained was purified using AMPure XP beads (Beckman Coulter, Cat. No. A63881). The purified cDNA was taken for multiplexed PCR amplification of the SARS-CoV-2 genome using ARTIC V3 (NEB) primer protocol (gives ∼400 bp amplicons) or the Midnight protocol from ONT (gives ∼1200 bp amplicons) The PCR amplicons were then barcoded by sequential End Repair/dA tailing (NEB reagents) and Native Barcode Ligation (ONT reagents). The samples were pooled, bead purified and adapter ligation (ONT reagents) was done with 200 ng of the barcoded pooled PCR amplicons. Nanopore sequencing of the library was carried out in sets of 12, 24, or 96 barcoded samples using the ONT R9.4.1 flow cells and GridION sequencer ((ONT). ONT library was also prepared according to ARTIC Midnight protocol PCR tiling of SARS-CoV-2 virus with rapid barcoding kit (SQK-RBK110.96) according to manufacturer’s protocol^41^.

### 4. SARS-CoV-2 whole genome sequencing using Illumina NextSeq550 platform (Illumina Platform)

Libraries were prepared using the defined protocol for Illumina Covid seq Ruo kits (Illumina Inc, USA). After annealing random hexamers, the first strand synthesis reaction was carried out on extracted RNA from Covid-19 positive samples. This was followed by cDNA amplification using a multiplex PCR protocol employing two primer pools-COVIDseq Primer Pool 1 and COVIDseq Primer Pool 2, containing a total of 98 amplicons to amplify the SARS-CoV-2 virus-specific sequences and 11 amplicons for amplifying human RNA. The PCR amplified products were further processed for Tag mentation and adapter ligation using 8 IDT for Illumina PCR Indexes Sets 1-4, each containing a total of 96 indices. Following enrichment and clean-up of tagmented amplicons as per recommended protocols, 96 sample batches were pooled together, with each batch containing a positive control (CPC HT) and one no template control (NTC). Pooled libraries were quantified using the Qubit 4.0 fluorometer (Invitrogen) and the Illumina Pooling calculator was used to combine all the pools to a final concentration of 4nM. For sequencing, the final pooled library was denatured, neutralized, and diluted to a final concentration of 1.4pM and paired-end sequencing was performed using the Illumina NextSeq 550 Sequencing Platform. Sequenced Read files were demultiplexed and converted to FASTQ using the FASTQ Generation Tool from Illumina BaseSpace. Reads were aligned to the reference Covid-19 genome (Wuhan-Hu-1, GenBank accession number MN908947.3) and variant calling was performed using the DRAGEN COVID Lineage application followed by lineage and clade analysis using Pangolin and Next-clade software ^42^.

### 5. Next-generation sequencing analysis of samples sequenced through the ONT platform

The MinKNOW 1.4.2 device control software embedded within the GridION sequencer was used to run the sequencer and collect the raw sequence data. The controlled parameters enable the user to analyze the raw fast5 files up to the variant calling. The Guppy base caller integrated within the MinKNOW 21.11.7 software was used for base calling and demultiplexing from the raw fast5. files using the base calling algorithms of Oxford Nanopore Technologies (https://community.nanoporetech.com) with a Phred quality cut-off score >7 on a GPU-Linux accelerated high-performance computing machine. Low-quality reads were filtered and discarded based on low Phred quality scores. From the amplicon sequences, a normalized read length of 300–500bp (amplicon size) is obtained which goes for demultiplexing using the ARTIC pipeline.

The demultiplexed fastq files were further analyzed downstream and aligned to the SARS-CoV-2 reference (MN908947.3) using the aligner Minimap2 v2.17^43^ through the command line execution which calls for the integration of Artic built-in pipeline Nextflow. The fastq files were indexed for variant calling from the Minimap output files using inbuilt medaka. Consensus fasta files are generated and used for further analysis for lineage identification.

### 6. Identification and assignment of lineages to sequences

Pangolin 4.0 software was used to assign the most likely lineage out of all currently designated lineages. It uses Scorpio to sanity-check specific lineages that correspond to variants. The consensus fasta sequences were realigned using minimap2 to the reference SARS-CoV-2 sequence to generate a fasta file with the non-coding regions masked out with N’s. The sequence is run through for QC check and pangoLearn with the help of VCF generates a lineage report. We ran 10456 fasta sequences to generate a lineage report ^44–46^.

### 7. Phylogenetic Analysis

Maximum likelihood phylogenetic trees were estimated using Nextclade Augur and rooted using Wuhan Hu-1 (MN908947.3) as an outgroup. Trees were constructed using the built-in IQ-Tree. Branch support statistics were generated using the ultrafast bootstrap method. Trees were visualized, explored, and labeled with associated metadata using Auspice to identify epidemiological links supported by the genomic data^47,48^.

### 8. Phylodynamics analysis and Molecular clock

For estimation of timeline and emergence and divergence of omicron variants, we used the Bayesian phylogenetic method using the Bayesian software package BEAST. For this analysis, we used the strict molecular clock model, the HKY + I + G nucleotide substitution model with an iteration of 3 million states each and sampling every 5000 steps. The convergence and the clade credibility of the trees we visualized using TreeAnnotator, ggtree, and ggplot ^49,50^.

### 9. Mutation profiling and Mutation analysis

SARS CoV-2 genome sequences for the Omicron variant, isolated between 15^th^ Dec 2021 to 10th Mar 2022 in South Africa (9484 sequences), Denmark (8979 sequences), Finland (6348 sequences), India (8478 sequences) were downloaded from the GISAID the global data science initiative database (https://gisaid.org/)^51,52^. For Pune, a total of 2540 Omicron sample sequences were considered for this analysis. The VCF files generated previously using Nextflow for mutation calling were used to extract the mutations and a heatmap was generated using “Complexheatmap” in R.

### 10. Cartographic analysis of lineage distribution

For making choropleth maps we obtained a GEOJSON shape file containing the boundaries of the wards of Pune. The ward office details were extracted from the address of each sample collected from the sample metadata. Using pandas, the geography information is extracted and mapped onto the geojson file in python, and the choropleth map was visualized using a plotly visualization tool. The maps were heat mapped based on the case counts for each month in each ward and a pie chart was generated to show lineage distribution in each ward.

## Data Availability

All data used in this study are available online at
GISAID Database

## Data Access

Data uploaded to GISAID

## Acknowledgments

Authors wish to thank the Pune Knowledge Cluster (PKC) for sample collection coordination. We also wish to thank Prof. Sanjeev Galande (IISER, Pune) for his help with the sequencing and Jacob John (CMC, Vellore) for guidance during sampling design.

## Ethics statement

The genome sequencing study was approved by the individual ethics committee of CSIR-NCL, Pune, and IISER Pune.

## Author contributions

AR wrote the manuscript. AR, KK, AG, JM, and LSS conceptualized the study and defined the sampling strategy. DN, SK, AK, and SN contributed to experimental design execution, data analysis, and implementation of software/code. All authors contributed to the experimental work. AR, KK, JM, AG, and DS contributed to the interpretation and critical revision of the manuscript. AR and LSS conceptualized the study. KK, DS, and AR supervised the research. LSS acquired the funding. PKC facilitated sample collection and distribution. RK supervised the dissemination of information to the policymakers. All authors contributed to the article and approved the submitted version.

## Funding

This work was supported in part by the Rockefeller Foundation, Council for Scientific and Industrial Research (CSIR), Villoo Poonawalla Foundation (VPF), Janakidevi Bajaj Gram Vikas Sanstha (JBGVS), Science and Engineering Board (SERB) Start-up Research Grant SRG/2020/000062, Department of Biotechnology (DBT)), RAD-22017/28/2020-KGD-DBT, INSACOG, and Johns Hopkins India Institute Covid Relief Fund.

## Conflict of Interest

The authors declare that they have no conflict of interest.

**Correspondence and requests for materials** should be addressed to AR

## Tables

**Supplemental Table 2:**
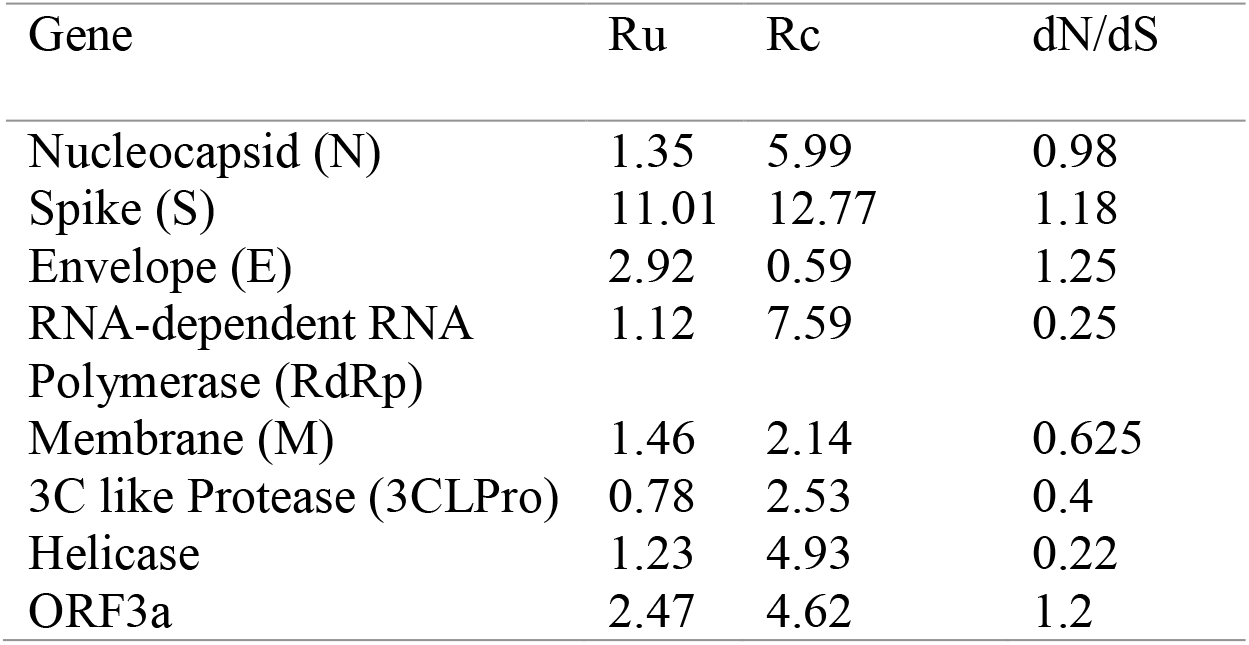
Gene-wise Mutation Rates and Evolutionary rates.

## Notes

### Competing Interest Statement

The authors have declared no competing interest.

### Funding Statement

This study was supported in part by the Rockefeller Foundation, Council for Scientific and Industrial Research (CSIR), Villoo Poonawalla Foundation (VPF), Janakidevi Bajaj Gram Vikas Sanstha (JBGVS), Science and Engineering Board (SERB) Start-up Research Grant SRG/2020/000062, Department of Biotechnology (DBT)), RAD-22017/28/2020-KGD-DBT, INSACOG, and Johns Hopkins India Institute Covid Relief Fund

### Author Declarations

The individual ethics committee of Council of Scientific and Industrial Research-National Chemical Laboratory of Pune and Indian Institutes of Science Education and Research of Pune gave ethical approval for this genome sequencing study

### Summary of Updates

The section containing the list of authors and their affiliations has been updated. Page numbers have been added. The materials and method section has been placed after the conclusion section.

