## Supplemental Table 2 for "A Tale of Two Waves: Diverse Genomic and Transmission Landscapes Over 15 Months of the COVID-19 Pandemic in Pune, India"

**Supplemental Table 2: Gene wise Mutation Rates and Evolutionary rates**

| Gene | Ru | Rc | dN/dS |
| --- | --- | --- | --- |
| Nucleocapsid (N) | 1.35 | 5.99 | 0.98 |
| Spike (S) | 11.01 | 12.77 | 1.18 |
| Envelope (E) | 2.92 | 0.59 | 1.25 |
| RNA dependent RNA Polymerase (RdRp) | 1.12 | 7.59 | 0.25 |
| Membrane (M) | 1.46 | 2.14 | 0.625 |
| 3C like Protease (3CLPro) | 0.78 | 2.53 | 0.4 |
| Helicase | 1.23 | 4.93 | 0.22 |
| ORF3a | 2.47 | 4.62 | 1.2 |
